# Age, Race, and Education as Moderators of Post-Stroke Cognitive Decline Following Dental Care

**DOI:** 10.1101/2025.07.21.25331931

**Authors:** Michael H. Parrish, Karly Pikel, Caitlin L. Scott, Haley N. VerKuilen, Souvik Sen

**Affiliations:** Department of Neurology, University of South Carolina School of Medicine, Columbia, SC

**Keywords:** post-stroke cognitive decline, dental care, age, race, education

## Abstract

Post-stroke cognitive decline (PSCD) poses a significant challenge to long-term recovery and quality of life following stroke, influenced by both fixed biological factors and modifiable health behaviors such as oral and dental care. In this data-driven exploratory analysis of the PREMIERS Phase II randomized trial (ClinicalTrials.gov NCT#02541032), we examined the moderating effects of clinical, biological, and demographic characteristics on the relationship between dental care and PSCD over a 12-month period. The study included 280 stroke/transient ischemic attack (TIA) survivors who received either intensive or standard dental care. Cognitive outcomes were assessed using the Montreal Cognitive Assessment (MoCA) at baseline and follow-up, with change in MoCA score as the primary outcome. Lasso regression was applied for empirically based feature selection of moderators, and bootstrapped multiple linear regression demonstrated that increased dental visits predicted relatively better cognitive outcomes in older adults (age interaction-term *β* = -0.664, *p* < 0.001), Black participants (race interaction-term *β* = -0.475, *p* < 0.05), and those with low-intermediate education levels (education interaction-term *β* = 0.413, *p* < 0.05). Exploratory graphs revealed that older adults, Black adults, and adults with low-intermediate education showed greater cognitive improvement with higher dental visit frequency, with the final model (including selected moderators) significantly predicting PSCD (*F*(11, 268) = 10.51, *p* = 5.17 x 10^−16^). These findings highlight the potential of equity-focused, precision-medicine interventions that incorporate dental care to mitigate PSCD in vulnerable stroke populations.

---

Post-stroke cognitive impairment (PSCI) and post-stroke cognitive decline (PSCD) represent major challenges to long-term recovery and quality of life following stroke. PSCI refers to cognitive deficits detected at a single post-stroke timepoint, while PSCD captures change in cognitive performance across multiple assessments. Both outcomes are clinically consequential: PSCI and PSCD - including progression to dementia - are associated with functional independence and quality of life (Quinn et al., 2018; El Husseini et al., 2023; Sadigh and Volovici, 2024), stroke recurrence (Moroney et al., 1997), and increased risk of mortality (Melkas et al., 2009; Oksala et al., 2009).

The cognitive consequences of stroke are shaped by complex interactions between biological injury and individual characteristics. Structural and vascular features such as lesion location, stroke subtype, and infarct volume contribute significantly to outcomes (Aam et al., 2020; Aamodt et al., 2023; Levine et al., 2025). For example, left hemisphere and posterior circulation strokes are particularly associated with cognitive deficit (Pendlebury and Rothwell, 2009; Siow et al., 2024), and medial temporal atrophy has differential effects depending on stroke laterality (Aamodt et al., 2022). These injury-related factors are typically fixed and not easily modifiable. However, demographic and clinical variables—such as age, sex, education, and comorbidities—also influence post-stroke cognitive trajectories and may provide avenues for targeted intervention (Rost et al., 2022; El Husseini et al., 2023).

In recent years, modifiable health behaviors and lifestyle factors have garnered attention for their potential to mitigate PSCI and PSCD. Physical activity, cognitive stimulation, sleep quality, and nutrition are increasingly studied for their protective roles (Obaid et al., 2020; Huang et al., 2022). Aerobic exercise, in particular, has shown consistent benefit for cognitive performance in stroke survivors (Moore et al., 2015; Steen Krawcyk et al., 2019), while treatment of sleep disorders like obstructive sleep apnea may also preserve cognition (Aaronson et al., 2016; Zhang et al., 2023). These insights have supported the development of multifactorial interventions, though effect sizes remain modest (Sun et al., 2021).

One emerging, yet underexplored, area for intervention is oral and dental health. Poor periodontal status may contribute to PSCD through systemic inflammation, microbiome dysregulation, and vascular dysfunction (Zhang et al., 2020; Wei et al., 2023; Shiraishi et al., 2024). Oral health is closely tied to immune aging (Ebersole et al., 2018) and may influence cognition through processes such as “inflammaging”—chronic, low-grade inflammation linked to aging and neurodegeneration (Franceschi et al., 2018; Finger et al., 2022). Modifying oral inflammation and health may thus offer a unique, scalable pathway to improve cognitive resilience, particularly in older adults.

We previously tested the effectiveness of dental health intervention improving stroke outcomes in the Periodontal Disease Treatment After Stroke or Transient Ischemic Attack (PREMIERS) Phase II randomized trial, which demonstrated that intensive periodontal treatment was safe and effective in improving vascular outcomes such as blood pressure and HDL levels (Sen et al., 2023). Our recent analysis of oral health data collected in PREMIERS showed that nearly half of stroke survivors with periodontal disease (PD) exhibited severe PSCI, and that severe PD independently associated with worse cognitive outcomes. Traditional risk factors such as Black race and greater stroke severity were also significant predictors, and advanced education was significantly protective (Pikel et al., 2025). In the present study, we conducted a secondary analysis of PREMIERS of cognitive health data to evaluate changes in cognition over a one-year period, with a focus on identifying subgroups (e.g., groups of different age, race, and education backgrounds) most likely to benefit from oral health care.

This analysis prioritized change-based assessment of PSCD, which more accurately captures stroke-related cognitive shifts than cross-sectional measures that may reflect preexisting pathology (El Husseini et al., 2023). Using a data-driven approach, we examined whether variables such as age, race, education, and sex moderated the relationship between dental care and PSCD. Understanding which populations benefit most could inform precision-based strategies for preventing PSCD in stroke survivors.

## Methods

### Study Population and Design

The PREMIERS clinical trial methods and results have been reported elsewhere (Sen et al., 2023; https://www.clinicaltrials.gov; Unique identifier: NCT 02541032), but the study will be summarized briefly here. PREMIERS was a multicenter phase II trial for which the primary objective was to evaluate the effect of intensive periodontal treatment on recurrent vascular events among stroke and high-risk TIA survivors. The sample of 280 patients was randomized in a 1:1 ratio to either intensive (n=140) or standard arm (n=140). Study activities were conducted in the clinical settings of the Neurology and Dentistry departments at both Prisma Health and The University of North Carolina at Chapel Hill. The study was approved by the Institutional Review Boards of both institutions. The results of this paper are based on a secondary analysis of the original study outcome data. This paper focuses on an analysis of all participants irrespective of treatment assignment, specifically examining the effects of number of dental visits attended on cognitive decline after the ischemic event.

### Periodontal Intervention

Full details of the periodontal intervention have been reported elsewhere (Sen et al., 2023), but a brief summary will be provided here. Intensive treatment involved supragingival and subgingival scaling and root planning using hand instruments and ultrasonic scalers under local anesthesia, extraction of teeth evaluated as being nonrestorable or too severely impacted by periodontitis, administration of local antibiotics, and provision of oral hygiene instructions. At baseline, intensive treatment participants were also given a Philips Sonicare ultrasonic toothbrush and an interdental cleaner with antibacterial mouth rinse. Standard treatment involved full-mouth supragingival scaling to remove only supragingival plaque and calculus and supragingival polishing with abrasive dental polishing paste. Standard treatment patients were also given education about their PD severity and referred to care if their oral condition needed immediate attention. Regardless of treatment assignment, all study participants were monitored for safety and progression of disease. At the end of the yearlong study period, the standard treatment group was offered the dental care involved in the intensive treatment.

### Cognitive Assessment

The Montreal Cognitive Assessment (MoCA) was administered at baseline and at the end of the 12-month study period. MoCA is an effective screening tool for PSCI and has been used in multiple studies of cognitive decline (Tan et al., 2017; Siqueira et al., 2019). The assessment evaluates overall cognitive capacity out of a total score of 30. It is based on evaluations on seven cognitive domains: visuospatial/executive, naming, attention, language, abstraction, recall, and orientation (Shi et al., 2018). MoCA may be a more sensitive measure of vascular cognitive impairment changes (compared to the Mini Mental State Examination, for example) because it includes components which test frontal-executive function and abstract reasoning (Tan et al., 2017). For our study, the PSCD score was calculated by subtracting the baseline score from 12-month follow-up score. Research has validated the MoCA for PSCD through comparison and association with formal neuropsychological test battery changes (Tan et al., 2017).

### Baseline Demographic, Clinical, and Brain Health Variables

Clinical and demographic information was obtained at the baseline visit before treatment began. Information was collected from the medical records about age/date of birth, sex, race, education level, hypertension, diabetes, substance use (smoking and alcohol), history of coronary artery disease, myocardial infarction, angina, coronary artery bypass graft, and atrial fibrillation (AFib). Level of education was operationalized as basic (1-12 years of education), intermediate (high school degree or equivalent), or advanced (completion of college degree or pursuing college-level education). Hypertension status was operationalized as a previous diagnosis of hypertension, irrespective of current medication treatment. Substance use was assessed through evaluation of current or past smoking cigarettes and current regular consumption of alcohol. AFib was operationalized as either past diagnosis or presentation on electrocardiogram (EKG) at admission. We assessed a measure of brain health by examining cerebral small vessel disease with the use of axial FLAIR imaging and the three-point Fazekas scale rating of white matter hyperintensities (WMH; lesions). MRI scans were acquired with a Siemens 1.5 T MRI scanner (Symphony Tim B17; Siemens Healthcare, Erlangen, Germany) or GE 1.5 T MRI Scanner (Discovery 450; MXR Imaging, San Diego, California, USA). Periventricular and deep WMH scores were rated separately (0-3) and then combined with a sum (0-6). Periventricular WMH (PVWMH) scores were rated in the following manner: 0 = absent; 1 = caps or pencil-thin lining; 2 = smooth “halo”; 3 = irregular periventricular signaling extending into the deep white matter. Deep WMH (DWMH) were rated in the following manner: 0 = absent, 1 = punctate foci; 2 = beginning of confluence of foci; 3 = large confluent areas of foci. TOAST subtype (Adams et al., 1993), stroke territory, and NIH Stroke Scale score were also evaluated and defined by the team’s vascular neurologist.

### Statistical Analyses

Statistical analyses were performed in R (R Core Team, 2023) and SPSS Version 29 (IBM Corp., 2022). First, descriptive statistics were analyzed for the candidate moderator variables: age, sex, race, BMI, NIHSS, income, hypertension status, diabetes status, coronary artery disease/myocardial infarction/coronary artery bypass graft, AFib, stroke territory, TOAST subtype, TIA status, history of smoking, current alcohol drinking status, and Fazekas WMH ratings. We specifically analyzed the distributions to examine any major outliers and visualized correlation plots to examine associations that could influence collinearity. We then completed missing variable cases with the multiple imputation approach implemented in the R package MICE (Multiple Imputation by Chained Equations). MICE uses iterative predictive models to fill in empty values by estimating relationships between variables. The MICE package creates multiple datasets for the missing values; to create a final dataset, we averaged missing values imputation values. To create interaction terms for each of the candidate variables, we first calculated z-scores for each candidate variable and multiplied these by the z-score for the number of study dental visits (labeled “Dental Visits”).

The data analysis was conducted in two steps: feature selection and final model estimation. In sum, we conducted a Lasso regression to automatically select the most important interaction terms in a data-driven fashion and then created a simplified multiple linear regression model for the purposes of interpretability and final significance testing. Lasso is a machine learning regression technique which involves introducing a L1 regularization penalty to the optimization function (James et al., 2023). This ultimately shrinks unimportant features to have a regression coefficient of zero. The non-zero regression coefficient terms are then used as the selected features for further analysis. Given the high number of interaction variables (n=17) and the drawbacks of alternatives such as stepwise linear regression (e.g., multiple testing issues), we decided to employ Lasso regression in our analyses. After feature selection, we conducted a final bootstrapped multiple linear regression analysis with the feature-selected interaction variables and their component terms (the components were input as predictors in the final model regardless of Lasso regression results). 95% confidence intervals generated from bootstrapping were then examined to evaluate final significance levels for all predictors of PSCD. Finally, we used bar chart graphs to evaluate the overall pattern of results producing significant interactions. Given the exploratory nature of the current research, we did not pursue further statistical comparison in mean levels of PSCD at specific levels of the interaction term component variables.

## Results

### Distributions of and Associations Between Candidate Moderator Variables

Figure 1 shows the variable distributions for the candidate moderators for the effect of Dental visits on PSCD. Notably, the distributions for Dental visits, BMI, Income, Baseline MoCA, and NIHSS were skewed; however, given that final analyses were conducted with bootstrapping methods, we were not concerned with the potential impact on results in the final simplified model. Supplemental Figure 1 shows the correlation matrix for the candidate variables. No variables were strongly correlated; therefore, multicollinearity was not expected to be an issue for regression analyses. While specific quantitative analyses and null hypothesis significance testing were not applied to these pairwise correlations, the strongest associations were found between: TIA and Territory, TIA and TOAST type, Race (Black) and Age, Race (Black) and Income, NIHSS and Baseline MoCA, Baseline MoCA and Income.

**Figure 1:**
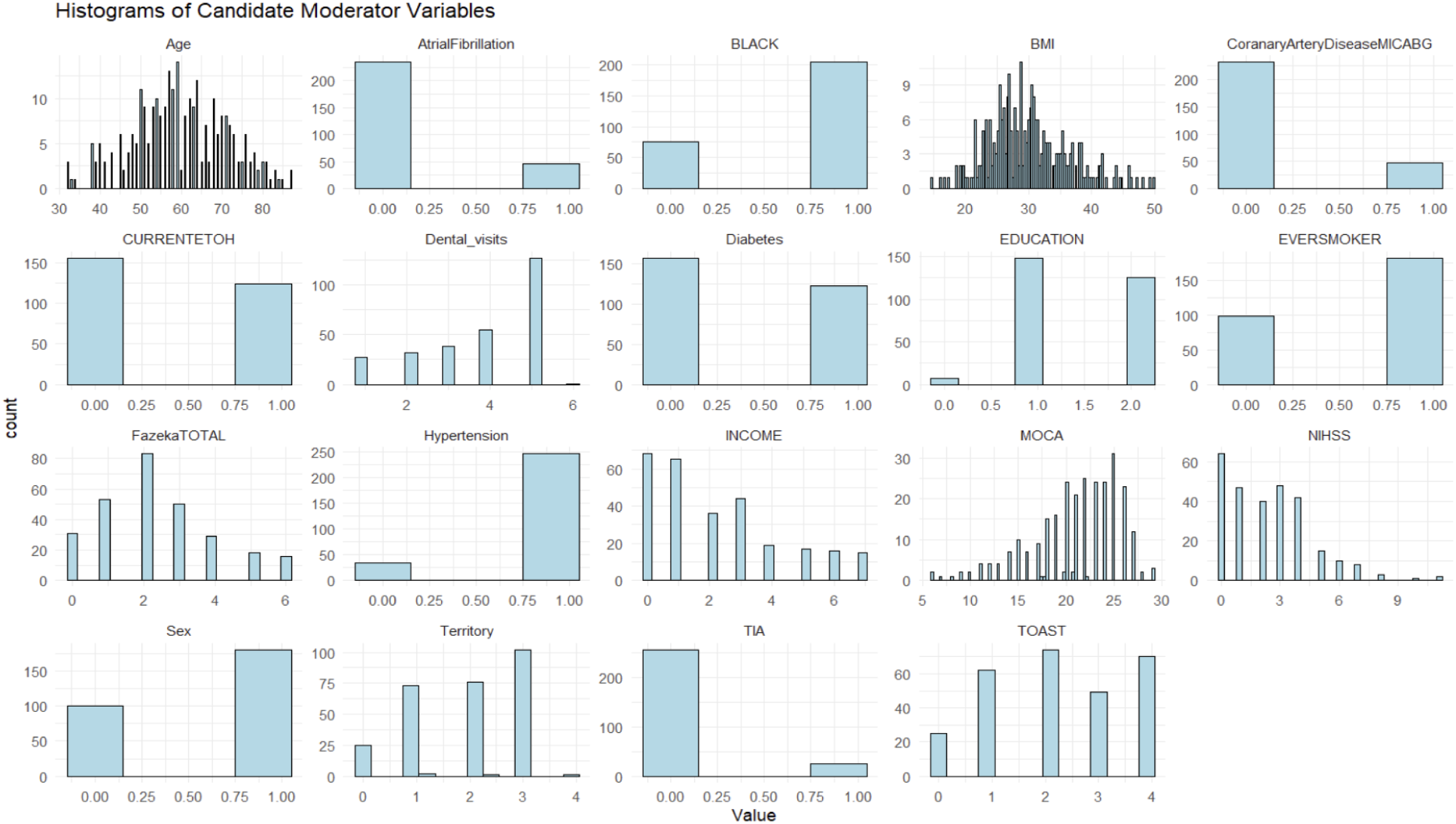
Histograms visualizing distributions of moderators tested in Lasso regression feature selection and multiple linear regression models.

### Feature Selection Results

Lasso regression analyses revealed that age-dental visits, race-dental visits, education-dental visits, and NIHSS-dental visits interaction variables were selected as significant features worth exploring in the context of multiple linear regression modeling. The other thirteen variables had their interaction term regression coefficients shrunk to zero given a lack of substantial relationship with the outcome variable PSCD. The order of the regularized weight’s magnitude/importance (descending order) was: race x dental visits (-.216), age x dental visits (-.214), education x dental visits (.096), and NIHSS x dental visits (-.008). In addition, Sex (Female) and Baseline MoCA variables were also selected based on Lasso regression results.

### Simplified Model Results

Multiple linear regression (MLR) analyses were then run with the interaction variables selected from feature selection and their component terms (e.g., both age and dental visits variables would be included because their interaction term was selected). The following variables were included: Dental visits (count), Sex (Female), Race (Black), Baseline MoCA, Age, Education, NIHSS, age x dental visits, race x dental visits, education x dental visits, and NIHSS x dental visits. The simplified model was found to be a significant prediction for PSCD (*F* (11, 268) = 10.51, *p* = 5.17 x 10^-16^). The predictors explained a significant amount of variance in the outcome: Multiple *R^2^* = 0.301, Adjusted *R*^2^ = 0.273. The following were found to be significant predictors in the simplified MLR model: Dental visits x Age (*β* = -0.664, *p* < 0.001; CI: [-1.067, -.240]), Dental visits x Race (Black) (*β* = -0.475, *p* = 0.023; CI: [-.903, -.059]), and Dental visits x Education (*β* = 0.413, *p* < 0.043; CI: [.035, .826]), Sex (Female) (*β* = -0.992, *p* = 0.018; CI: [-1.807, -.170]), Baseline MoCA (*β* = -0.439, *p* < 0.001; CI: [-.570, -.314]).

### Graphical Exploration of Age, Race, and Education Interaction Effects

Figures 2-4 depict exploratory visualizations of the relationship between significant moderators, number of dental visits, and PSCD. Following statistical analysis, we next interrogated the three significant interaction effects by visually inspecting the specific relationships between different levels of the moderators and predictors and the outcome variable PSCD. For the bar graph depicting the significant Dental visit x Age interaction, we found that increasing number of dental visits was uniquely beneficial for older adults, specifically those in their seventies. This benefit was not similarly demonstrated for adults younger than 70 years old. For the bar graph depicting the significant Dental visit x Race (Black) interaction, we found that higher number of dental visits was uniquely beneficial for Black adults. Surprisingly, non-Black participants showed increased levels of cognitive impairment at the highest numbers of dental visits. For the bar graph depicting the significant Dental visit x Education interaction, we found that only adults with intermediate levels of education (high school graduate, no college degree or higher) benefitted from the increased number of dental visits. However, it should be noted only seven participants reported basic level of education. These analyses may suffer from a lack of statistical power reflected in the results. In summary, the post-stroke cognition of older adults, Black adults, and adults with low-intermediate levels of education appeared to benefit most from increasing effects of more dental treatment.

**Figure 2:**
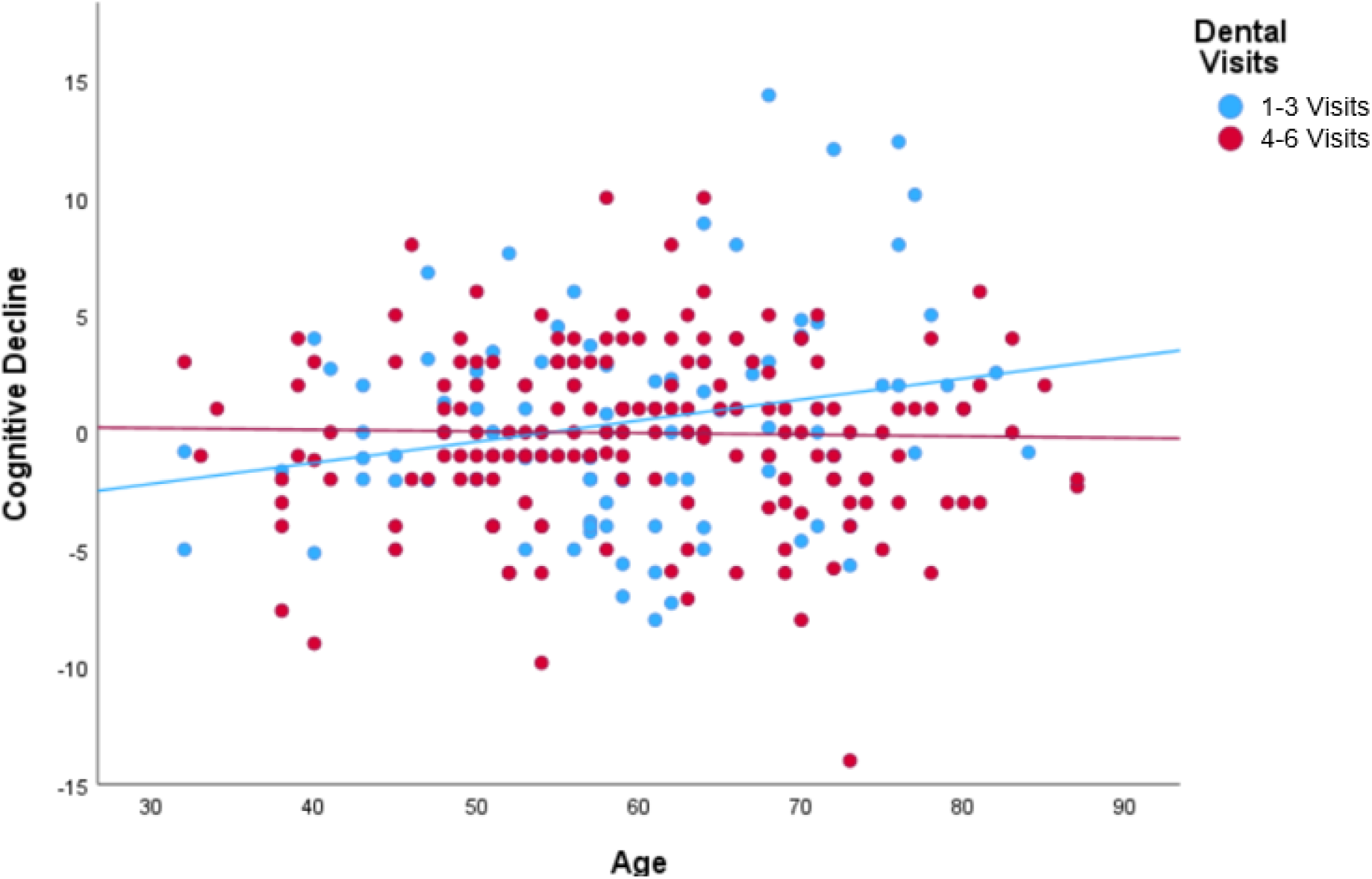
Scatterplot visualizing interaction between number of dental visits and age influencing post-stroke cognitive decline.

**Figure 3:**
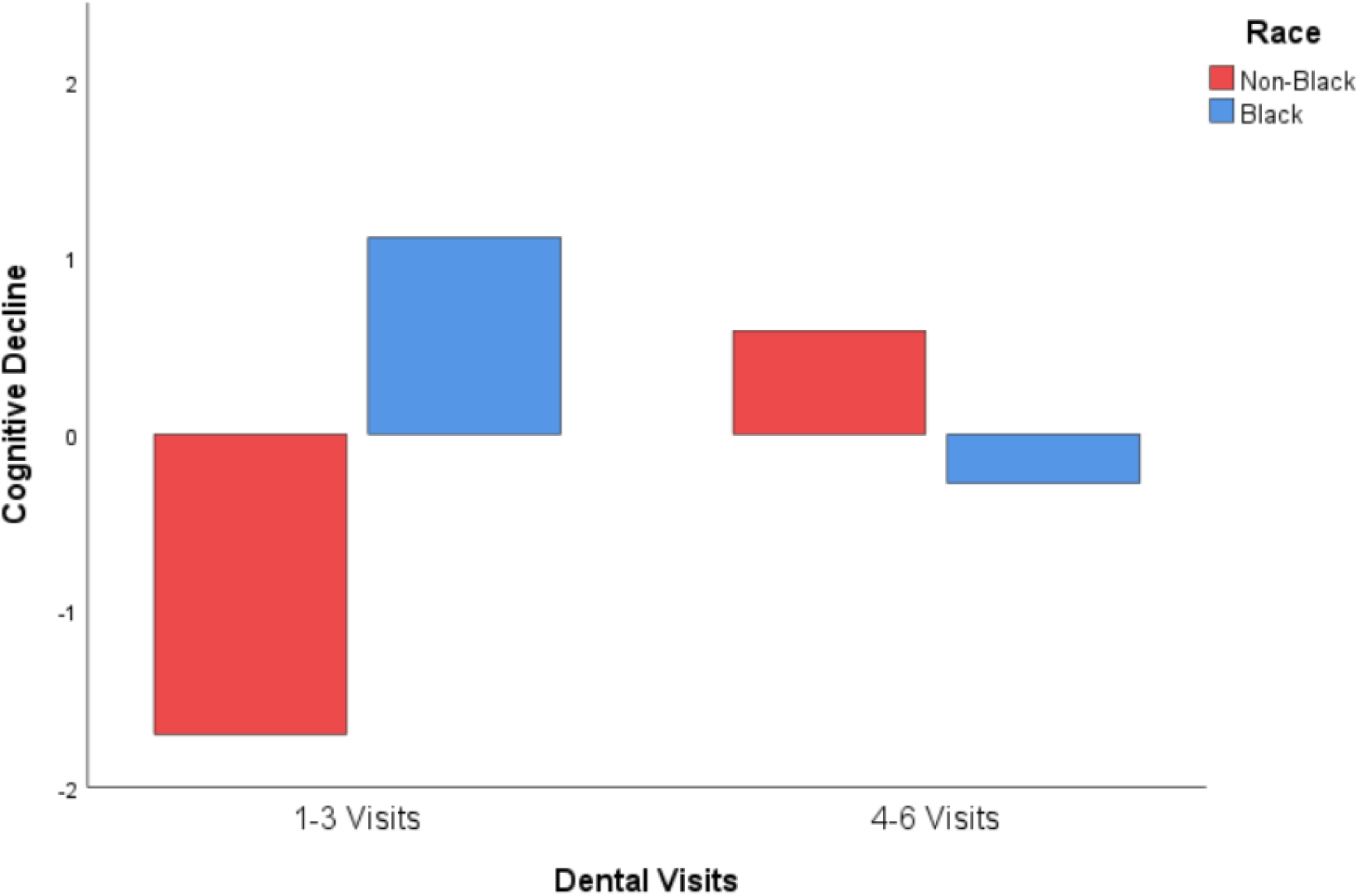
Bar chart visualizing interaction between number of dental visits and race influencing PSCD.

**Figure 4:**
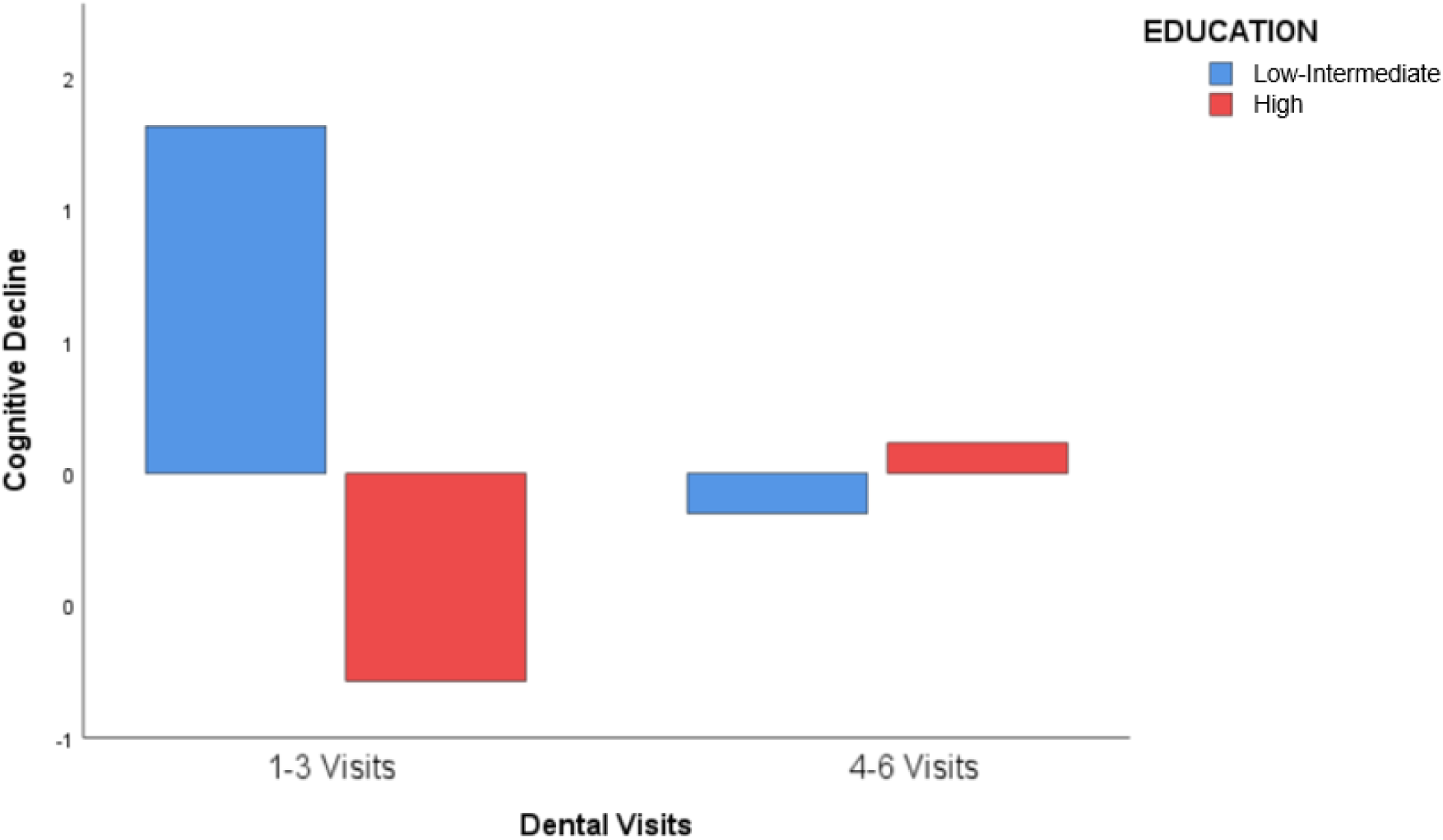
Bar chart visualizing interaction between number of dental visits and education influencing PSCD.

## Discussion

In this secondary analysis of the PREMIERS trial, we applied data-driven statistical approaches to identify three key demographic moderators—age, race, and education—that shaped the relationship between dental care and PSCD. Specifically, greater exposure to dental treatment was associated with improved cognitive outcomes among older adults, Black adults, and individuals with low to intermediate levels of education.

Age emerged as the most reliably significant moderator, with older individuals deriving the greatest cognitive benefit. This is consistent with the robust relationship between aging and vulnerability to PSCD (Hua et al., 2022; Lo et al., 2022). Age-related changes in brain structure and immune function—including diminished white matter integrity, vascular remodeling deficits, and altered inflammatory profiles—may predispose older adults to steeper cognitive decline (Cabeza et al., 2018; Rost et al., 2022; Aamodt et al., 2023). However, these same biological vulnerabilities may also enhance responsiveness to interventions that modulate systemic inflammation. Oral health interventions may help reverse components of “inflammaging” and restore immune resilience (Franceschi et al., 2018; Finger et al., 2022). Aging is also linked to reduced functional connectivity in neural networks supporting psychological resilience (Chen et al., 2025), and it is plausible that health-promoting behaviors like oral care can strengthen these networks via improved self-efficacy and engagement (Chen et al., 2025). These mechanisms suggest that aging may not only be a risk factor but also a point of leverage for targeting cognitive outcomes in stroke survivors.

Race was also a significant moderator, with Black adults experiencing greater cognitive benefit from dental treatment. This aligns with research showing that Black individuals often experience more severe PD, and greater susceptibility to PSCD (Sarfo et al., 2020; Gillone et al., 2023; Wang et al., 2023). This observed pattern may reflect underlying inflammatory burden, genetic predisposition, and differential intervention response. Despite these disparities, behavioral interventions—such as computerized cognitive training have shown greater positive effects in Black adults (Nwosu et al., 2024). Similarly, vascular interventions like hypertension management, offer a unique opportunity to mitigate racial cognitive disparities. For example, one cohort study found racial disparities in cognition between Black and White adults were in part explained by levels of systolic blood pressure (the previously observed disparity in cognition was no longer statistically significant after controlling for systolic blood pressure; Levine et al., 2020). Given the systemic link between PD and vascular conditions such as hypertension, targeted dental interventions may yield the same effects. Our findings suggest that oral care may represent a similarly effective strategy for reducing cognitive disparities by targeting modifiable inflammatory and vascular pathways.

Perhaps most notably, educational attainment moderated cognitive outcomes in an unexpected direction: individuals with low-intermediate education, compared to high education (i.e., high school diploma but no college degree) experienced the greatest benefit from dental care. This contrasts with the assumption that higher education always confers greater cognitive resilience. Some evidence suggests that more highly educated individuals may harbor greater latent neuropathology that is masked by compensatory mechanisms until a threshold is passed—after which rapid cognitive decline ensues (Scarmeas et al., 2006; Verlinden et al., 2016). In stroke populations, executive function decline post-event has been found to be steeper in college-educated individuals (Springer et al., 2025). These individuals may also be less responsive to health interventions if their neuropathological burden is too severe to be no longer modifiable. By contrast, individuals with intermediate education may represent a “sweet spot” of cognitive reserve—where moderate pathology remains responsive to systemic interventions like oral care. This interpretation is consistent with findings from stroke cohorts in Korea and Europe, where lower education predicted worse recovery, but also suggested greater capacity for treatment-related gains (Shin et al., 2020; Elgh and Hu, 2023).

Additionally, it is noteworthy for future research focused on dental care intervention effects that we found baseline cognition and female sex were predictive of PSCD, however neither moderated the effect of dental visits. Higher baseline global cognition was related to lower levels of PSCD, potentially indicating the individuals with higher levels of cognitive reserve were more resistant to PSCD, although this has yet to be tested comprehensively (Tao et al., 2023; Li et al., 2022). Additionally, our results show that female sex, compared to male sex, related to decreased PSCD. While current overall research on sex-specific patterns of PSCD is mixed, this finding is in line with at least some evidence that women may be uniquely protected from neurological deficits in certain domains of cognition, such as verbal memory (Zinman et al., 2023; Exalto et al., 2023). Though these variables may not directly interact with dental care to influence cognitive health, they should be included in statistical models predicting PSCD to account for their independent effects.

Taken together, these findings suggest that interventions to prevent PSCD may be more effective when tailored to subgroups based on age, race, and educational background. Most existing cognitive rehabilitation strategies yield small to moderate effects (Sun et al., 2021), and few are designed with population-specific responsiveness in mind. Our results highlight the need for stratified trial designs, targeted recruitment strategies, and subgroup analyses to maximize and detect efficacy in vulnerable groups (Elkind et al., 2020; Chavez et al., 2024).

Despite the strengths of this study, several limitations should be acknowledged. The analysis was underpowered to detect small moderation effects, and findings related to race were essentially limited to comparisons between Black and White participants. Furthermore, all results were produced from a relatively small sample size and need to be interpreted with caution. Additionally, we did not measure depressive symptoms and depression may have confounded or mediated the statistical relationships between age, sex, education, dental care, and PSCD (Lo et al., 2022; Shin et al., 2022). Finally, cognitive outcomes were assessed only over one year; longer-term follow-up is needed to determine the durability of treatment effects.

In conclusion, this study identifies three subgroups—older adults, Black adults, and individuals with low-intermediate education—who may uniquely benefit from dental care as a strategy to mitigate PSCD. These findings support the integration of oral health into broader cognitive intervention frameworks and emphasize the need for equity-focused, precision approaches in stroke rehabilitation.

## Data Availability

All data produced in the present study are available upon valid request to the authors.

---

**Supplemental Figure 1:**
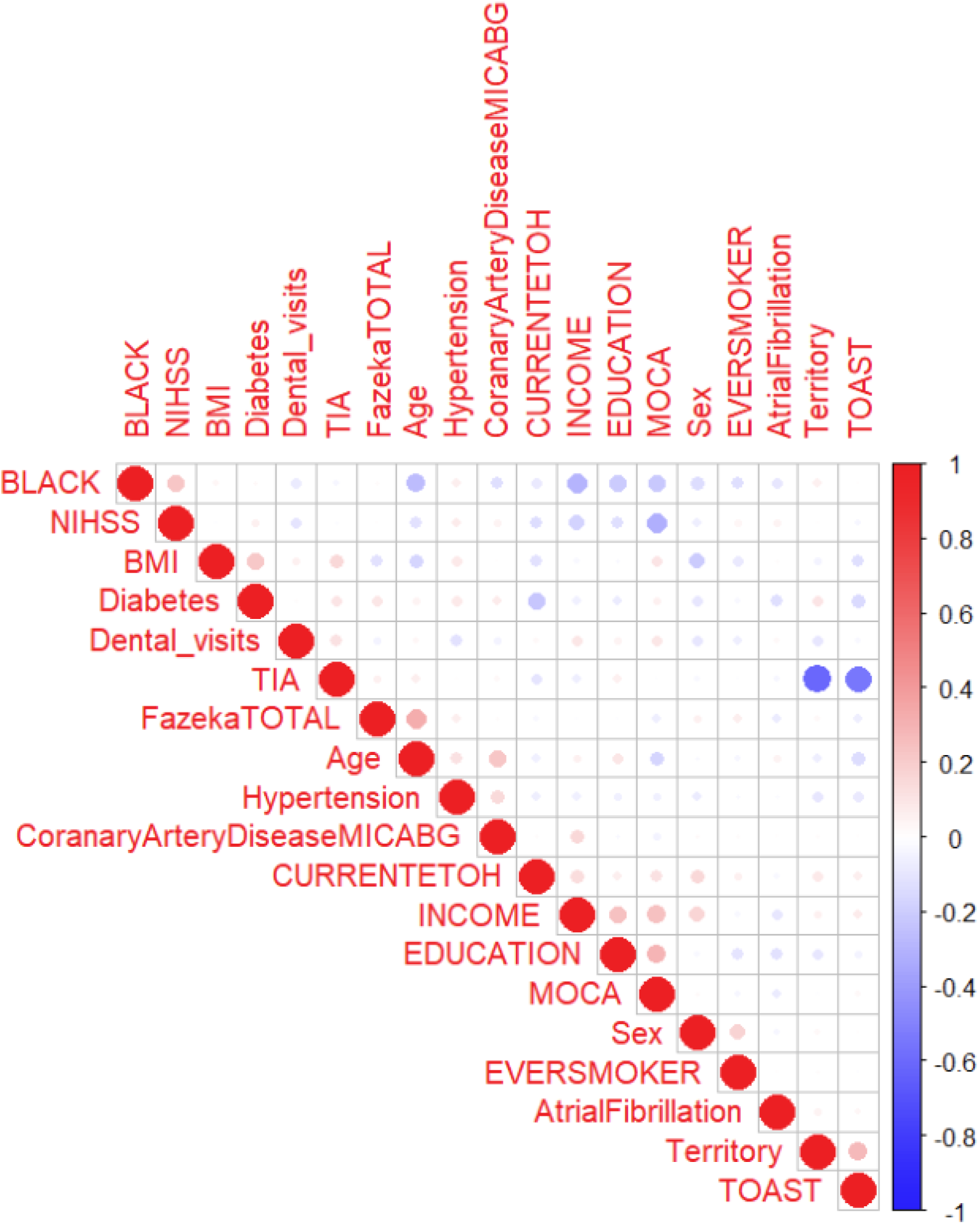
Correlation matrix depicting bivariate relationships between candidate moderator variables.

